# Underutilization of Lipoprotein(a) Testing and Its Association with Coronary Interventions: PITT-LIPID Real-world Registry Insights

**DOI:** 10.64898/2026.09.03.26361336

**Authors:** Romana Awan, Hongtian Wang, Floyd Thoma, Jianhui Zhu, Jacqueline Levene, Suresh Mulukutla, Agnes Koczo, Anum Saeed

**Affiliations:** Department of Internal Medicine, Mercy Catholic Medical Center, Darby, PA; Heart and Vascular Institute, UPMC, Pittsburgh, PA; Providence Heart Clinic - Hollywood, Portland, OR; Center for Cardiovascular Disease Prevention Research, University of Pittsburgh School of Medicine, Pittsburgh, PA

**Author notes:** Corresponding Author: Anum Saeed, MD, 200 Lothrop St, BST 1721, Pittsburgh, PA 15213 E, Twitter: @AnumSaeedMD.

**Keywords:** Lipoprotein (a), Primary prevention, coronary artery disease, atherosclerotic cardiovascular disease, coronary revascularization, ASCVD

## Abstract

**Background:** Lipoprotein(a) [Lp(a)], is leading risk factor for premature atherosclerotic cardiovascular disease. Multi-societal guidelines recommend to test Lp(a) amongst adults at least once, however, real-world testing data in primary prevention populations remains scarce.

**Objectives:** To investigate the utilization of Lp(a) testing in a primary prevention cohort and associated outcomes.

**Methods:** We conducted a retrospective cohort study using data from the PITT-LIPID registry, an electronic health record (EHR) based database of primary prevention patients (ages 20-79 years) across a tristate healthcare network. Lp(a) testing status was assessed, and those with Lp(a) measurements were stratified by Lp(a) levels ≤125 versus >125 nmol/L. Multivariable Cox proportional hazards models were used to assess associations between Lp(a) levels and coronary artery interventions [percutaneous coronary intervention (PCI), coronary artery bypass grafting (CABG)], and mortality.

**Results:** Among 687,780 patients without known coronary artery disease, only 6,340 (0.9%) patients underwent Lp(a) testing. Compared with untested patients, those tested for Lp(a) had higher low density lipoprotein cholesterol, greater percentages of statin and aspirin use, and lower prevalence of diabetes and smoking. Patients with Lp(a) testing had higher rates of PCI (4.5% vs 1.4%; p-value <0.001) and CABG (1.4% vs 0.4%; p-value <0.001) but lower all-cause mortality (2.0% vs 5.0%; p-value < 0.001) incidence. Lp(a) >125 nmol/L was independently associated with risk of PCI (HR 1.67, 95% CI [1.16-2.41]), p=0.006.

**Conclusion:** Lp(a) testing was underutilized and associated with higher burden of PCI events in a primary prevention population across a contemporary healthcare network with diverse practice types.

## 1. Introduction

Despite recent advances in prevention and therapeutic options, atherosclerotic cardiovascular disease (ASCVD) is a leading cause of morbidity and mortality worldwide (1). Even with optimal lipid-lowering therapy, substantial residual risk for ASCVD events persists (2,3). Within the past decade, lipoprotein(a) [Lp(a)] has emerged as one of the primary causes of premature ASCVD and associated morbidity and mortality (4,5).

Despite the recently updated recommendations for Lp(a) testing once in a lifetime for all adults (6,7) universal Lp(a) screening is not routinely implemented in the United States. Although no direct Lp(a) modifying agents which have proven cardiovascular outcomes benefit are commercially available – several pharmacological treatments are currently under investigation. Importantly, observational data suggest that use of aspirin and statins may confer benefit in individuals with high Lp(a) (8,9). Data on use of these preventive therapies specifically in patients with elevated Lp(a) to mitigate Lp(a)-associated ASCVD risk is not well characterized in contemporary clinical practices (5,10).

To address this gap, we investigated real-world patterns of Lp(a) testing, utilization of commercially available primary prevention therapies and long-term risk of ASCVD as well as mortality events within a large, healthcare system–wide registry of patients without previously established coronary artery disease.

## 2. Methods

### 2.1 Population cohort

We conducted a retrospective cohort study using data from the PITT-LIPID registry, an electronic health record (EHR) based database in a large academic healthcare network details of which have been published (11,12). Briefly, the PITT-LIPID registry cohort was amassed using an EHR data from University of Pittsburgh Medical Center (UPMC) health system – a network of private practices and academically affiliated clinical practices across the Western Pennsylvania, New York and Maryland areas including over 40 hospitals and 400 outpatient clinics.

This retrospective cohort study was approved by the Institutional Review Board (IRB) of UPMC and UPMC Quality Improvement Committee. Given the use of de-identified data from existing medical records, IRB waived the requirement for informed consent.

The primary prevention cohort for the current study included a group of patients aged 20 to 79 years with at least 2 UPMC health care interactions at a UPMC facility between January 2013 and December 2025 with at least one lipid profile result within 180 days of the first health care system interaction (index visit). The date of indexed UPMC facility visit served as the baseline date of entry to the cohort. All included patients had at least one baseline covariate dataset available in the system for review and follow up visits for inclusion in the registry.

### 2.2 Covariates-Derived risk

Baseline characteristics of the study population were defined using data within 180 days of the index date of entry into the cohort. These included sociodemographic data (age, sex, and race/ethnicity, low-density lipoprotein cholesterol, and the remaining PREVENT equation (13) variables including; total cholesterol, high-density lipoprotein cholesterol, treatment for hypertension, systolic blood pressure, current smoking status, and diabetes. Demographic variables, such as age (difference between the index date and date of birth), race (categorized as Black or White), sex, and medication use were derived from administrative data sources within the UPMC Clinical Data Warehouse.

Diabetes was defined as the use of a diabetes medication before the baseline date plus either ICD-9 and ICD-10 diagnosis codes. Blood pressure treatment was defined as an active prescription on the baseline date for at least 1 of the following medication classes, taken as a single agent or in combination formulations: diuretics, angiotensin-converting enzyme inhibitors, angiotensin II receptor inhibitors, α-blockers, β-blockers, and calcium channel blockers. Smoking history was categorized as current or not current (a composite of never and former smoking).

For subgroup analysis, patients with at least one lipoprotein(a) [Lp(a)] measurement were included, and they were further stratified based on Lp(a) levels (≤125 vs >125 nmol/L). Patients with extreme Lp(a) values (>400 nmol/L) were excluded to minimize outliers. Individuals with missing variables required for analysis were excluded.

Baseline demographics, comorbidities, lipid profile and associated medication use (aspirin and statin) were compared between groups.

### 2.3 Outcomes

The primary outcomes included incident coronary artery disease including myocardial infarction and revascularization, incident transient ischemic attacks and ischemic events, heart failure hospitalization, incident ASCVD and mortality, per risk category.

Incident ASCVD events were defined as a composite outcome of ischemic stroke, myocardial infarction, and revascularization with percutaneous coronary intervention and/or coronary artery bypass graft. All outcomes were surveyed and included as defined per the ICD9 and ICD10 procedural coding in EHR.

Mortality was assessed using the United States Social Security Death Index. Our health care system is exempt from the 3-year delay period by the Social Security Administration.

### 2.4 Statistical Analysis

Baseline descriptive statistics of the study sample were initially analyzed to detect outliers and missing values. Among all predictor variables, only Low-density lipoprotein cholesterol [LDL-C] and triglycerides contained missing values. The proportion of missing values for these two variables was less than 5% of the total study population. Therefore, participants with missing LDL-C or triglyceride measurements were excluded from the analysis. Descriptive characteristics were compared among groups with and without Lp(a) testing, and presented as mean and SD for continuous variables, and frequencies and proportions for categorical variables. The difference of mean across the ASCVD groups was assessed by Student’s t-tests, while the difference of categorical frequencies is compared using the χ^2^ test.

Multivariable Cox proportional hazards regression models were used to assess associations between Lp(a) levels and the composite outcomes. Hazard ratios (HR) with 95% confidence intervals (CI) were reported, and statistical significance was defined as p-value <0.05. The proportional hazards assumption was assessed using Schoenfeld residuals, and no violations were detected.

All statistical analyses were performed using R (version 4.4.2; R Core Team, Vienna, Austria).

## 3. Results

### 3.1 Baseline demographics

A total of 687,780 primary prevention patients were included in analysis, of which only 0.9% (n=6340) had at least one Lp(a) test done (**Table 1a**). Patients undergoing Lp(a) testing were older (mean age 51.3 ± 12.3 vs. 49.9 ± 14.3 years, *p*<0.001), more likely to self-identify as White (89.8%) as compared to those without Lp(a) testing. Those with Lp(a) tested also had significantly lower prevalence of diabetes (6.6% vs. 10%, *p*<0.001) and smoking (10.4% vs. 19.1%, *p*<0.001).

**Table 1a:** Baseline Characteristics of Patients in PITT-LIPID Registry Database Comparing Those with and without Lp(a) Testing.

|  | Overall (n = 687,780) | With Lp(a) tests (n = 6340) | Without Lp(a) test (n = 681440) | p-value |
| --- | --- | --- | --- | --- |
| **Demographics** |  |  |  |  |
| Age (year) | 49.9±14.3 | 51.3±12.3 | 49.9±14.3 | <0.001 |
| Female | 396139 (57.6%) | 3618 (57.1%) | 392521 (57.6%) | 0.400 |
| White | 605727 (88.1%) | 5693 (89.8%) | 600034 (88.1%) | <0.001 |
| Black/African American | 52101 (7.6%) | 353 (5.6%) | 51748 (7.6%) | <0.001 |
| Current Smoking | 131152 (19.1%) | 661 (10.4%) | 130491 (19.1%) | <0.001 |
| SBP (mm Hg) | 126.4±15.9 | 125.0±15.1 | 126.5±15.9 | <0.001 |
| BMI | 30.3±7.3 | 28.7±6.2 | 30.3±7.3 | <0.001 |
| Diabetes | 68373 (9.9%) | 417 (6.6%) | 67956 (10.0%) | <0.001 |
| **Medications** |  |  |  |  |
| Aspirin | 101705 (14.8%) | 1289 (20.3%) | 100416 (14.7%) | <0.001 |
| Statins | 123081 (18.0%) | 1530 (24.1%) | 121551 (17.8%) | <0.001 |
| PCSK9 | 101 (0.014%) | 10 (0.16%) | 86 (0.013%) | <0.001 |
| **Laboratory Results** |  |  |  |  |
| Total Cholesterol (mg/dL) | 193.5±36.1 | 205.3±39.5 | 193.4±36.1 | <0.001 |
| HDL Cholesterol (mg/dL) | 52.9±15.2 | 54.8±15.7 | 52.9±15.2 | <0.001 |
| LDL Cholesterol<br>(mg/dL) | 115.2±33.0 | 125.4±35.5 | 115.1±32.9 | <0.001 |
| Triglycerides<br>(mg/dL) | 130.1±76.6 | 127.9±75.3 | 130.2±77.2 | 0.020 |

Abbreviations: Lp(a), lipoprotein(a) SBP, systolic blood pressure; BMI, body mass index; PCSK9, proprotein convertase subtilisin/kexin type 9 inhibitor; HDL, high-density lipoprotein; LDL, low-density lipoprotein

**Table 1b:** Baseline Characteristics of Patients with at Least One Lipoprotein (a) Testing.

| | With Lp(a)<br>tests*<br>(n = 6255) | Lp(a) level ≤ 125 (n =<br>4610) | Lp(a) level ><br>125<br>(n = 3034) | p-value |
| --- | --- | --- | --- | --- |
| **Demographics** |  |  |  |  |
| Age (year) | 51.3±12.4 | 51.3±12.5 | 51.3±12.1 | 0.970 |
| Female | 3562 (56.9%) | 2559 (55.5%) | 1003 (61.0%) | 0.001 |
| White | 5620 (89.8%) | 4169 (90.4%) | 1451 (88.2%) | 0.012 |
| Black/African American | 342 (5.5%) | 193 (4.2%) | 149 (9.1%) | <0.001 |
| Current Smoking | 645 (10.3%) | 467 (10.1%) | 178 (10.8%) | 0.457 |
| SBP (mm Hg) | 125.0±15.1 | 125.0±15.0 | 124.9±15.4 | 0.746 |
| BMI | 28.7±6.2 | 28.7±6.2 | 28.8±6.1 | 0.700 |
| **Medications** |  |  |  |  |
| Statins | 859 (13.7%) | 615 (13.3%) | 244 (14.8%) | 0.142 |
| **Comorbidities** |  |  |  |  |
| Diabetes | 412 (6.6%) | 297 (6.4%) | 115 (7.0%) | 0.477 |
| **Laboratory Results** |  |  |  |  |
| Total Cholesterol<br>(mg/dL) | 205.1±39.4 | 203.4±39.1 | 209.7±39.9 | <0.001 |
| HDL Cholesterol<br>(mg/dL) | 54.8±15.8 | 54.6±15.8 | 55.5±15.5 | 0.048 |
| LDL Cholesterol<br>(mg/dL) | 125.1±35.4 | 123.4±35.0 | 129.9±35.8 | <0.001 |
| Triglycerides (mg/dL) | 127.8±75.5 | 129.0±78.2 | 124.3±67.2 | 0.018 |

\*Total n after removal of all Lp(a) values > 400 nmol/L
Abbreviations: SBP, systolic blood pressure; BMI, body mass index; HDL, high-density lipoprotein; LDL, low-density lipoprotein

Among serological variables studied, patients who received Lp(a) testing had higher LDL-C (125.4 vs. 115.1 mg/dL, *p*<0.001) and total cholesterol (205.3 vs 193.4 mg/dL, p<0.001). There was a higher percentage of Lp(a) tested patients on statins (24.1% vs. 17.8%, *p*<0.001) or aspirin (20.3% vs. 14.7%, *p*<0.001). Interestingly, BMI was lower in the Lp(a) tested group (28.7 vs. 30.3 kg/m², *p*<0.001). (**Table 1a**)

Among patients with at least one Lp(a) measurement; those with Lp(a) >125 nmol/L (n=3,034) had no significant difference in age (average age 51.3 years in both groups), were more commonly females than those with Lp(a) ≤125 nmol/L (61.0% vs. 55.5%, *p* = 0.001) (**Table 1b**). Further, those with Lp(a) >125 nmol/L were more likely to self-identify as White (88.2% vs. 90.9%, *p* <0.05). There were no significant differences in smoking status (10.8% vs. 10.1%, *p* = 0.457) or systolic blood pressure (124.9 vs. 125 mmHg, *p* = 0.746) between 2 Lp(a) groups. BMI also did not differ significantly between groups (28.8 vs. 28.7 kg/m², *p* = 0.70). Statin use was similar (14.8% vs. 13.3%, *p* = 0.142), as was the prevalence of diabetes (7.0% vs 6.4%, *p* = 0.477).

Lipid panel results showed significantly higher total cholesterol (209.7 vs. 203.4 mg/dL, *p* < 0.001), LDL-C (129.9 vs. 123.3 mg/dL, *p* < 0.001) in patients with Lp(a) >125.

### 3.2 Statin Use

Table 2 shows the patients who underwent Lp(a) testing and were on statin therapy at the time of ASCVD events. Amongst a total of n=1590 patients, a higher percentage were on moderate-intensity statins as compared to patients without Lp(a) testing (moderate-intensity statin: 73.77% vs 76.77%, p=0.005). In contrast, among the 124,577 patients without Lp(a) testing, 11.56% (n=14,403) were on high-intensity, 76.77% (n=95,640) on moderate-intensity, and 6.99% (n=8,705) on low-intensity statins. Use of multiple statin doses was more frequent in the Lp(a) tested group (6.48%) compared to the non-tested group (3.48%). Less than 1.5% of the cohort reported being on pitavastatin or “unknown statin” category and were not used in further analysis.

**Table 2:** Statin Intensity Use Amongst Primary Prevention Patients Stratified by Lp(a) Testing.

|  | With Lp(a) tests<br>(n= 1590) | Without Lp(a) test<br>(N = 124577) | P-value |
| --- | --- | --- | --- |
| High-intensity Statin | 209 (13.14%) | 14403 (11.56%) | 0.055 |
| Moderate-intensity Statin | 1173 (73.77%) | 95640 (76.77%) | 0.005 |
| Low-intensity Statin | 84 (5.28%) | 8705 (6.99%) | 0.009 |
| Multiple Doses | 103 (6.48%) | 4332 (3.48%) | <0.001 |

### 3.3 Aspirin and statin use with Lp(a) stratification ≤125 vs >125 nmol/L

Subgroup stratification by Lp(a) levels (≤125 vs. >125nmol/L) revealed notable differences in medication use patterns. (table 3) In both subgroups, a great majority of patients were not receiving either statins or aspirin (66.5% in the Lp(a) ≤125 nmol/L group and 64.9% in the >125 group).

**Table 3:** Proportions of Patients Who Had Statin or Aspirin Use Stratified by Lp(a) levels.

| Total Patients<br>(n=6255) | Lp(a) ≤ 125 nmol/L<br>N = 3946 | Lp(a) > 125 nmol/L<br>N = 2309 | P-value |
| --- | --- | --- | --- |
| Statin only | 517(13.10%) | 342(14.81%) | 0.063 |
| Aspirin only | 409(10.36%) | 218(9.44%) | 0.258 |
| Both Statin and Aspirin | 397(10.06%) | 251(10.87%) | 0.332 |
| None | 2623(66.47%) | 1498(64.88%) | 0.209 |

\*Total n after removal of all Lp(a) values > 400 nmol/L

Statin only use nominally more common in the Lp(a) >125 nmol/L level group (14.8%) compared to the ≤125 group (13.1%), (*p* = 0.063). Similarly, therapy with both statin and aspirin was essentially the same among patients with >125 nmol/L Lp(a) (10.9% vs. 10.1%, *p* = 0.332). Conversely, aspirin only use was more frequent in Lp(a) group ≤ 125 (10.4% vs 9.4%,) in Lp(a) > 125 group, however not statistically significant (p=0.258).

### 3.4 Cardiovascular Interventions and Mortality

Among total of 687,780 patients in study population, the proportion of patients who underwent PCI was significantly higher in the Lp(a) tested group (4.5%) compared to the non-Lp(a) group (1.4%); p-value <0.001 (**Table 4a**). Similarly, CABG procedure occurred more frequently in the Lp(a) tested group (1.4%) than in those without Lp(a) test (0.4%); p <0.001. In contrast, all-cause mortality was lower in the Lp(a) tested group (2.0%) compared to the non-Lp(a) group (5.0%), with a significant p-value <0.001.

**Table 4a:**
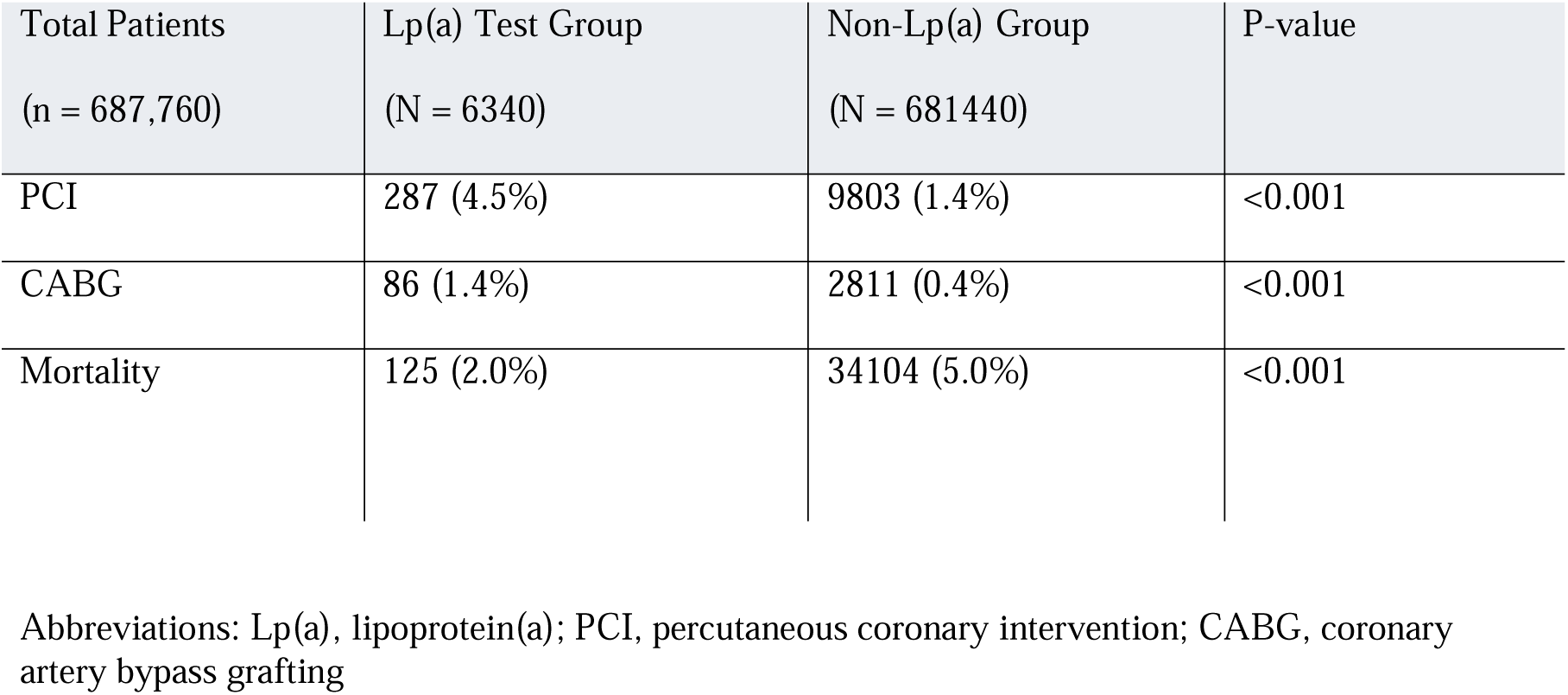
Proportion of coronary artery intervention and all-cause mortality events in patients with and without lipoprotein (a) Testing.

**Table 4b:** Association between Lipoprotein(a) levels and Coronary Artery Interventions and Mortality.

| Clinical Outcome | Lp(a) comparison<br>(nmol/L) | Adjusted<br>Hazard Ratio | 95% CI | p-value |
| --- | --- | --- | --- | --- |
| PCI | >125 vs ≤ 125 | 1.67 | 1.16-2.41 | 0.006 |
| CABG | >125 vs ≤125 | 2.02 | 0.98-4.17 | 0.056 |
| Mortality | 75-124 vs <75 | 0.59 | 0.28-1.23 | 0.161 |
|  | 125-174 vs <75 | 0.92 | 0.47-1.80 | 0.809 |
|  | ≥175 vs <75 | 0.44 | 0.22-0.87 | 0.018 |

Abbreviations: Lp(a), lipoprotein(a); CI, Confidence Interval; PCI, percutaneous coronary intervention; CABG, coronary artery bypass grafting

### 3.5 Lp(a) levels and association with coronary artery interventions (PCI, CABG) and Mortality

In comparison to Lp(a) levels ≤ 125, Lp(a) >125nmol/L was significantly associated with an elevated risk of undergoing PCI (HR□=□1.67, 95% CI [1.16–2.41], *p*□=□0.006), independent of other cardiovascular risk factors and LDL-C levels (**Table 4b)**. Patients with Lp(a)≥175 nmol/L had significantly reduced risk of mortality compared to those with Lp(a) <75□(HR□=□0.44, 95% CI [0.22–0.87], *p*□=□0.018).

**Supplemental Tables S** shows predictors of PCI, CABG and mortality amongst this population. Additional predictors of PCI (**Table S1**) included male sex in comparison to female sex (HR□=□2.09, 95% CI [1.58-2.76], *p*□<□0.001), higher systolic blood pressure (HR= 1.02, 95% CI [1.01-1.03], *p* < 0.001), diabetes (HR = 1.80, 95% CI [1.23-2.62], *p* = 0.002), and low HDL levels. In contrast, being a current non-smoker compared to non-smoker (HR = 0.51, 95% CI [0.37-0.70], *p* < 0.001) and higher HDL-C levels (≥40mg/dL: HR = 0.58, 95% CI [0.43-0.79], *p* < 0.001) were protective.

## 4. Discussion

In this large, contemporary health-system registry of 687,780 primary prevention patients, we report three principal findings. First, lipoprotein(a) testing was profoundly undertested, performed in fewer than 1% of patients despite guideline recognition of Lp(a) as a clinically relevant risk enhancer.

Second, testing was not applied systematically but was instead concentrated among patients who already carried a recognizable dyslipidemia signature - those with higher total and LDL cholesterol-rather than being ordered on the basis of universal, guideline-directed criteria.

Third, consistent with prior epidemiological data, elevated Lp(a) was independently associated with increased risk of subsequent coronary revascularization, particularly PCI.

This study’s results have clinical and public health policy implications; to our knowledge, this is among the largest characterizations of real-world Lp(a) testing behavior in a dedicated primary prevention population - the very group in whom universal, one-time screening is intended to deliver its greatest value. Universal screening earns its rationale by identifying elevated Lp(a) before a first ASCVD event, when primary prevention can still alter the trajectory. Documentation of how testing actually behaves in this population is essential, and yet a risk factor with a validated causal link to ASCVD is being measured in fewer than 1% of eligible adults across a large contemporary healthcare system. Although universal testing of Lp(a) was recently introduced in the American College of Cardiology/American Heart Association Dyslipidemia Management Guidelines 2026, but the recommendation was not new; the National Lipid Association had already endorsed one-time universal measurement in its 2019 scientific statement and reaffirmed it in its mid-2024 update (6,7). Our work demonstrates that this marked underutilization of lipoprotein(a) testing prevails almost 18 months after the NLA recommendations were published. That a decade of accumulating guidance has produced such low testing rates tells us that recommendation alone does not change behavior.

Further, the pattern of who does get the testing makes this a sharper problem. The observed demographic and clinical differences between tested and non-tested populations in our data suggest that Lp(a) measurement is being applied selectively with preference among patients with higher total and LDL cholesterol as well as smoking but lower diabetes. Selective testing of this kind guarantees that the patients whose risk is silently elevated by Lp(a), absent any other lipid abnormality, are precisely the ones who go unmeasured. This is the opposite of what universal screening is meant to accomplish.

While prior reports have demonstrated limited uptake of Lp(a) testing and highlighted the need for more systematic incorporation into routine cardiovascular risk assessment (14,15); most of these data have come from secondary prevention populations which are at a high risk of recurrent events (16,17). Testing gaps in that setting, while important, understate the problem. Failure to screen is most consequential precisely in primary prevention, where an elevated Lp(a) may be the only signal of risk and where the window to intervene before an event is still open.

Finally, an important observation in this study is the coexistence of higher rates of coronary interventions with lower mortality among patients who underwent Lp(a) testing. This apparent paradox is likely reflective of selection and treatment effects rather than a protective effect of the testing itself. Patients undergoing Lp(a) testing may represent a subgroup with greater healthcare access, increased access to specialty care, and more surveillance, leading to earlier detection of coronary disease and timely revascularization. In clinical practice, identification of elevated Lp(a) may also prompt escalation of preventive therapies, contributing to improved observed outcomes (14,15).

## 5. Conclusion

In a large contemporary U.S. healthcare system registry, Lp(a) testing was rare and selectively applied. Elevated Lp(a) was independently associated with increased risk of coronary revascularization, yet its identification did not consistently prompt treatment intensification - a fundamental translational gap between what guidelines recognize and what clinical practice delivers. As targeted Lp(a)-lowering therapies move toward the clinic, closing this gap is no longer optional: without systematic testing, the patients these therapies are designed to help will remain invisible until they present with disease.

## 6. Limitations

This study has several limitations. First, it is a single center registry, generalizability to more diverse populations may be limited. Although previously published literature shows similarly low Lp(a) testing across contemporary healthcare systems across the US. Second, the retrospective observational design limits the ability to establish causality, and findings may be influenced by confounders despite thoroughly adjusted models in analyses. Third, Lp(a) testing was performed in a very small proportion of the cohort (0.9%), introducing significant selection bias, as patients undergoing testing likely represent a higher-risk and more closely monitored subgroup.

## Data Availability

All data produced in the present study are available upon reasonable request to the authors

## 7. Acknowledgments

None.

## Abbreviations

ASCVD: atherosclerotic cardiovascular disease
CABG: coronary artery bypass grafting
CI: confidence intervals
EHR: electronic health record
HR: Hazard ratios
IRB: Institutional Review Board
LDL-C: Low-density lipoprotein cholesterol
Lp(a): lipoprotein(a)
PCI: percutaneous coronary intervention
UPMC: University of Pittsburgh Medical Center

