## Supplemental Table S1, S2, S3 for "Underutilization of Lipoprotein(a) Testing and Its Association with Coronary Interventions: PITT-LIPID Real-world Registry Insights"

**Supplemental Table S1: Cox Proportional Hazards Regression Model on PCI for Patients Who Had At Least 1 Lp(a) Test**

| N = 6255  After removing all Lp(a) values > 400 | Hazard Ratio | 95% Confidence Interval | P value |
| --- | --- | --- | --- |
| Age | 1.04 | (1.02, 1.05) | <0.001 |
| Male (ref: Female) | 2.09 | (1.58, 2.76) | <0.001 |
| Race (ref: Black/African American) | | | |
| White | 0.85 | (0.54, 1.34) | 0.481 |
| Other | 1.03 | (0.48, 2.24) | 0.940 |
| Currently Not Smoking | 0.51 | (0.37, 0.70) | <0.001 |
| SBP | 1.02 | (1.01, 1.03) | <0.001 |
| BMI | 1.00 | (0.98, 1.03) | 0.733 |
| Diabetes History (ref: none) | 1.80 | (1.23, 2.62) | 0.002 |
| *LDL* Cholesterol *(ref: <70 mg/dL)* | | | |
| 70–99 mg/dL | 1.44 | (0.65, 3.16) | 0.370 |
| 100–129 mg/dL | 1.11 | (0.50, 2.46) | 0.800 |
| 130–159 mg/dL | 1.49 | (0.67, 3.33) | 0.327 |
| 160–189 mg/dL | 1.63 | (0.70, 3.80) | 0.255 |
| ≥190 mg/dL | 1.68 | (0.66, 4.31) | 0.279 |
| *HDL* Cholesterol *(ref: <40 mg/dL)* | | | |
| 40–59 mg/dL | 0.58 | (0.43, 0.79) | <0.001 |
| ≥60 mg/dL | 0.47 | (0.31, 0.71) | <0.001 |
| *Triglycerides (ref: <150 mg/dL)* | | | |
| 150–199 mg/dL | 1.28 | (0.93, 1.77) | 0.135 |
| 200–249 mg/dL | 1.08 | (0.69, 1.69) | 0.741 |
| 250–499 mg/dL | 0.84 | (0.51, 1.40) | 0.507 |
| ≥500 mg/dL | 1.67 | (0.40, 6.98) | 0.485 |
| Current Medication (ref: None) | | | |
| Aspirin only | 0.88 | (0.54, 1.45) | 0.620 |
| Statin only | 1.13 | (0.73, 1.74) | 0.593 |
| Both statin and aspirin | 0.91 | (0.57, 1.45) | 0.681 |
| *Lp(a) (ref:* ≤ *125 nmol/L)* | | | |
| >125 nmol/L | 1.67 | (1.16, 2.41) | 0.006 |
| *Interactions between Current Medicine and Lp(a) Level (ref: None and Lp(a)* ≤ *125* | | | |
| Aspirin only: Lp(a) > 125 | 1.76 | (0.86, 3.63) | 0.123 |
| Statin only: Lp(a) > 125 | 0.73 | (0.34, 1.53) | 0.400 |
| Both: Lp(a) > 125 | 1.20 | (0.59, 2.45) | 0.618 |

**Abbreviations:** Lp(a), lipoprotein(a) SBP, systolic blood pressure; BMI, body mass index; LDL Cholesterol, low-density lipoprotein Cholesterol; HDL Cholesterol, high-density lipoprotein Cholesterol. P < 0.05 is significant.

**Supplemental Table S2: Cox Proportional Hazards Regression Model on CABG for Patients Who Had At Least 1 Lp(a) Test**

| N = 6255  After removing all Lp(a) values > 400 | Hazard Ratio | 95% Confidence Interval | P-value |
| --- | --- | --- | --- |
| Age | 1.05 | (1.02, 1.07) | <0.001 |
| Male (ref: Female) | 2.57 | (1.49, 4.44) | <0.001 |
| Race (ref: Black/African American) | | | |
| White | 0.66 | (0.31, 1.40) | 0.277 |
| Other | 2.57 | (0.88, 7.55) | 0.085 |
| Currently Not Smoking | 0.56 | (0.31, 1.00) | 0.052 |
| SBP | 1.03 | (1.01, 1.04) | <0.001 |
| BMI | 1.01 | (0.97, 1.05) | 0.691 |
| Diabetes History (ref: none) | 1.40 | (0.73, 2.68) | 0.311 |
| *LDL* Cholesterol *(ref: <70 mg/dL)* | | | |
| 70–99 mg/dL | 0.64 | (0.18, 2.30) | 0.497 |
| 100–129 mg/dL | 1.22 | (0.35, 4.17) | 0.757 |
| 130–159 mg/dL | 1.85 | (0.53, 6.49) | 0.337 |
| 160–189 mg/dL | 1.46 | (0.37, 5.81) | 0.595 |
| ≥190 mg/dL | 0.42 | (0.04, 4.22) | 0.465 |
| *HDL* Cholesterol *(ref: <40 mg/dL)* | | | |
| 40–59 mg/dL | 0.28 | (0.17, 0.48) | <0.001 |
| ≥60 mg/dL | 0.21 | (0.10, 0.48) | <0.001 |
| *Triglycerides (ref: <150 mg/dL)* | | | |
| 150–199 mg/dL | 0.65 | (0.32, 1.29) | 0.215 |
| 200–249 mg/dL | 1.46 | (0.77, 2.80) | 0.250 |
| 250–499 mg/dL | 0.84 | (0.37, 1.88) | 0.665 |
| ≥500 mg/dL | 1.51 | (0.20, 11.66) | 0.694 |
| *Current Medication (ref: None)* | | | |
| Aspirin only | 1.12 | (0.44, 2.85) | 0.806 |
| Statin only | 1.85 | (0.82, 4.18) | 0.139 |
| Both statin and aspirin | 2.81 | (1.34, 5.89) | 0.006 |
| *Lp(a) (ref:* ≤*125 nmol/L)* | | | |
| >125 nmol/L | 2.02 | (0.98, 4.17) | 0.056 |
| *Interactions between Current Medicine and Lp(a) Level (ref: None and Lp(a)* ≤ *125* | | | |
| Aspirin only: Lp(a) > 125 | 0.87 | (0.20, 3.78) | 0.856 |
| Statin only: Lp(a) > 125 | 0.57 | (0.14, 2.29) | 0.426 |
| Both: Lp(a) > 125 | 0.83 | (0.27, 2.56) | 0.749 |

**Abbreviations:** Lp(a), lipoprotein(a) SBP, systolic blood pressure; BMI, body mass index; LDL Cholesterol, low-density lipoprotein Cholesterol; HDL Cholesterol, high-density lipoprotein Cholesterol. P < 0.05 is significant.

**Supplemental Table S3: Cox Proportional Hazards Regression Model on Mortality for Patients Who Had At Least 1 Lp(a) Test without Interaction Terms**

| N = 6255  After removing all Lp(a) values > 400 | Hazard Ratio | 95% Confidence Interval | P-value |
| --- | --- | --- | --- |
| Age | 1.11 | (1.08, 1.13) | <0.001 |
| Male (ref: Female) | 1.03 | (0.67, 1.57) | 0.902 |
| Race (ref: Black/African American) | | | |
| White | 0.45 | (0.23, 0.87) | 0.018 |
| Other | 0.81 | (0.27, 2.45) | 0.706 |
| Currently Not Smoking | 0.59 | (0.34, 1.02) | 0.058 |
| SBP | 1.01 | (1.00, 1.02) | 0.131 |
| BMI | 1.03 | (0.99, 1.06) | 0.154 |
| Diabetes History (ref: none) | 2.00 | (1.21, 3.32) | 0.007 |
| *LDL* Cholesterol *(ref: <70 mg/dL)* | | | |
| 70-99 mg/dL | 2.28 | (0.68, 7.59) | 0.180 |
| 100-129 mg/dL | 2.18 | (0.64, 7.42) | 0.212 |
| 130-159 mg/dL | 2.36 | (0.67, 8.23) | 0.179 |
| 160-189 mg/dL | 2.93 | (0.78, 10.99) | 0.112 |
| >190 mg/dL | 4.25 | (1.04, 17.30) | 0.044 |
| *HDL* Cholesterol *(ref: <40 mg/dL)* | | | |
| 40-59 mg/dL | 0.43 | (0.26, 0.71) | <0.001 |
| ≥60 mg/dL | 0.54 | (0.30, 0.98) | 0.044 |
| *Triglycerides (ref: <150 mg/dL)* | | | |
| 150-199 mg/dL | 0.78 | (0.46, 1.34) | 0.372 |
| 200-249 mg/dL | 0.86 | (0.42, 1.74) | 0.668 |
| >250 mg/dL | 0.88 | (0.42, 1.82) | 0.726 |
| *Current Medicine (ref: None)* | | | |
| Aspirin only | 1.02 | (0.58, 1.80) | 0.935 |
| Statin only | 1.13 | (0.66, 1.97) | 0.653 |
| Both Statin and Aspirin | 1.34 | (0.79, 2.30) | 0.280 |
| *Lp(a) (ref: < 75 nmol/L)* | | | |
| 75-124 nmol/L | 0.59 | (0.28, 1.23) | 0.161 |
| 125-174 nmol/L | 0.92 | (0.47, 1.80) | 0.809 |
| ≥175 nmol/L | 0.44 | (0.22, 0.87) | 0.018 |

**Abbreviations:** Lp(a), lipoprotein(a) SBP, systolic blood pressure; BMI, body mass index; LDL Cholesterol, low-density lipoprotein Cholesterol; HDL Cholesterol, high-density lipoprotein Cholesterol. P < 0.05 is significant.
